# Predicting and forecasting the impact of local outbreaks of COVID-19: Use of SEIR-D quantitative epidemiological modelling for healthcare demand and capacity

**DOI:** 10.1101/2020.07.29.20164566

**Authors:** Eduard Campillo-Funollet, James Van Yperen, Phil Allman, Michael Bell, Warren Beresford, Jacqueline Clay, Matthew Dorey, Graham Evans, Kate Gilchrist, Anjum Memon, Gurprit Pannu, Ryan Walkley, Mark Watson, Anotida Madzvamuse

## Abstract

**Background:** The world is experiencing local/regional hot-spots and spikes of the severe acute respiratory syndrome coronavirus 2 (SARS-CoV-2), which causes COVID-19 disease. We aimed to formulate an applicable epidemiological model to accurately predict and forecast the impact of local outbreaks of COVID-19 to guide the local healthcare demand and capacity, policy making, and public health decisions.

**Methods:** The model utilised the aggregated daily COVID-19 situation reports (including counts of daily admissions, discharges, and bed occupancy) from the local NHS hospitals and COVID-19 related weekly deaths in hospitals and other settings in Sussex (population 1·7M), Southeast England. These datasets corresponded to the first wave of COVID-19 infections from 24 March to 15 June 2020. A novel epidemiological predictive and forecasting model was then derived based on the local/regional surveillance data. Through a rigorous inverse parameter inference approach, the model parameters were estimated by fitting the model to the data in an optimal sense and then subsequently validated.

**Results:** The inferred parameters were physically reasonable and matched up to the widely used parameter values derived from the national datasets.^28^ We validate the predictive power of our model by using a subset of the available data and compare the model predictions for the next 10, 20, and 30 days. The model exhibits a high accuracy in the prediction, even when using only as few as 20 data points for the fitting.

**Conclusions:** We have demonstrated that by using local/regional data, our predictive and forecasting model can be utilised to guide the local healthcare demand and capacity, policy making, and public health decisions to mitigate the impact of COVID-19 on the local population. Understanding how future COVID-19 spikes/waves could possibly affect the regional populations empowers us to ensure the timely commissioning and organisation of services. The flexibility of timings in the model, in combination with other early warning systems, produces a timeframe for these services to prepare and isolate capacity for likely and potential demand within regional hospitals. The model also allows local authorities to plan potential mortuary capacity and understand the burden on crematoria and burial services. The model algorithms have been integrated into a web-based multi-institutional toolkit, which can be used by NHS hospitals, local authorities, and public health departments in other regions of the UK and elsewhere. The parameters, which are locally informed, form the basis of predicting and forecasting exercises accounting for different scenarios and impact of COVID-19 transmission.

## Introduction

Since SARS-CoV-2 was identified in December 2019,^1^ COVID-19 has swiftly and rapidly spread to nearly all countries in the world, becoming an ongoing global world pandemic that has required unprecedented international, national, and regional interventions to try and contain its spread.^1,2^ Unlike the 1918-19 H1N1 pandemic, which is considered one of the greatest medical disasters of the 20th century,^1^ the spread of COVID-19 has unfolded live on multimedia platforms with real-time updates, statistics and with remarkable reporting accuracy,^3^ and yet reliable, accurate, and data-validated epidemiological modelling with forecasting and prediction capabilities remains largely out of reach.^1,4–8^ Given the lack of widely accessible pharmaceutical interventions, such as vaccination and antiviral drugs, epidemiological modelling has been thrust to the forefront of world organisations’ and governments’ responses, rapid decision-making, and public health interventions and policy.^1,4,9–11^ Until these pharmaceutical interventions become widely available, the only measures for infection prevention and control are self or group-isolation (quarantine), testing and contact tracing, physical distancing, decontamination, use of personal protective equipment, wearing masks and hygiene measures. A lot of these unprecedented actions/decisions have resulted in complete lockdowns of countries and economies, and yet these decisions are based on qualitative/quantitative predictions/models using national datasets outside the countries imposing the lockdowns on the basis of these models. A fair criticism of the underlying approach has been the lack of rigorous model validation and applicability given the datasets available at the time of the study, the lack of risk assessment associated with the decisions and their impact on the healthcare demand, capacity, and delivery and subsequently the lack of precision forecasting that is driven by data.^8,12^ Unfortunately, early epidemiological models needed to make assumptions out of necessity about parameters and disease progression. Therefore, given the lack of data at the early stages of the pandemic, the predictions of these models were near impossible to validate.^1,4,6,7,9^ At the forefront of these epidemiological models that have played a pivotal role in guiding national public health policy and healthcare responses that include the current social distancing, contact tracing, and quarantine measures, is the well documented Imperial College London model.^1^ The societal and economic impact of the aforementioned decisions have hardly been quantified, only estimates in the range of trillions of dollars loss to the world economy are reported.^13,14^ A few models dealing with decision-making within the COVID-19 crisis have been reported;^9,15,16^ however, these lack the power of model prediction and forecasting based on appropriate variables and datasets.

In order to understand the temporal dynamics of COVID-19, a lot of modelling work has been undertaken, focusing primarily on national datasets from China, Italy, Spain, UK, and the USA.^1,4,6,7,9,17–20^ Given the inhomogeneous nature of such datasets, accurate predictions and forecasting of the spread of COVID-19 is challenging.^8,21^ Where such predictions were made, caveats accompanied these predictions simply because of the lack of rigorous mathematical and statistical validation of the models and the lack of robust data on which mathematical assumptions are based.^1,4,6–9,19,20^ Forecasting requires ample historical information/datasets, which were lacking during the first wave of COVID-19. Current state-of-the-art forecasting models are based, on the one hand, on time series analysis without an underlying dynamic epidemiological model.^6,8,22,23^ On the other hand, where forecasting is based on epidemiological models,^6,24^ these lack rigorous validation, sensitivity analysis, and analysis with respect to identifiability of parameters and therefore have limited forecasting power. An interesting approach is proposed in Bertozzi et al. (2020)^4^ where three models were presented depending on the forecasting timescales; an exponential growth model, a self-exciting branching process, and the classical susceptible-infected-recovered (SIR) compartmental model. The exponential growth model is assumed valid at the early stages of the pandemic, the self-exciting branching process models the individual count data going into the development of the pandemic, and the SIR is a macroscopic mean-field model that describes the pandemic dynamics as it approaches the peak of the infection and disease. Another interesting and alternative approach is to build machine learning and artificial intelligence techniques on top of epidemiological models to allow for model predictions and forecasting.^6^ This approach, so far, has been applied to national datasets from the USA but no regional modelling of this type has been undertaken.

The use of local datasets is critical for managing and mitigating COVID-19 secondary spikes/waves and re-infection within local communities.^25^ Already there is ample evidence that local forecasting models could help local/regional authorities to plan lockdowns, restrictions, opening of schools/universities, as well as planning for healthcare demand and capacity. For example, during the summer of 2020 all the 50 states in the USA started to relax lockdown restrictions, however, several states soon after either put on hold their efforts to open fully or started to backtrack due to the resurgence of COVID-19 infections and the start of secondary waves.^8^ At the same time in the UK, cities such as Leicester, Bradford, and Oldham were in the midst of experiencing secondary COVID-19 waves and re-infection. Similarly, in Australia, the city of Melbourne in the state of Victoria was in stage 4 lockdown while the remainder of the state was in stage 3 lockdown.^10,11^ During the first wave, Australia was hailed as a global success story in suppressing the spread of COVID-19 and even at the height of the initial outbreak, it only reported a little over 600 infections a day. A similar story emerged in Spain with regions in Catalonia undergoing secondary lockdowns. The usefulness of national models, in all these countries, is not clear in terms of being able to predict and forecast the emergence of such spikes, waves or new incidences locally until they have already taken place, which is too late. We propose therefore an alternative quantitative predictive approach which gives local (and national) authorities the ability to predict and forecast COVID-19 scenarios based on their current historical datasets to visualise future dynamic temporal trends of the infection/disease progression for healthcare planning purposes.

In this study, we want to demonstrate the usefulness and utility of a locally data-driven epidemiological model, based on recent datasets from the three adjoining regions in Sussex, Southeast England (i.e. Brighton and Hove City Council, East and West Sussex County Councils), to make predictions and forecast to guide local/regional decision-making and healthcare delivery. The approach is based on a modified SIR-type model, (Figure 1), that has been formulated to reflect the dynamics of the combined Sussex populations of approximately 1·7 million and the mathematical interpretation of the data available.

**Figure 1:**
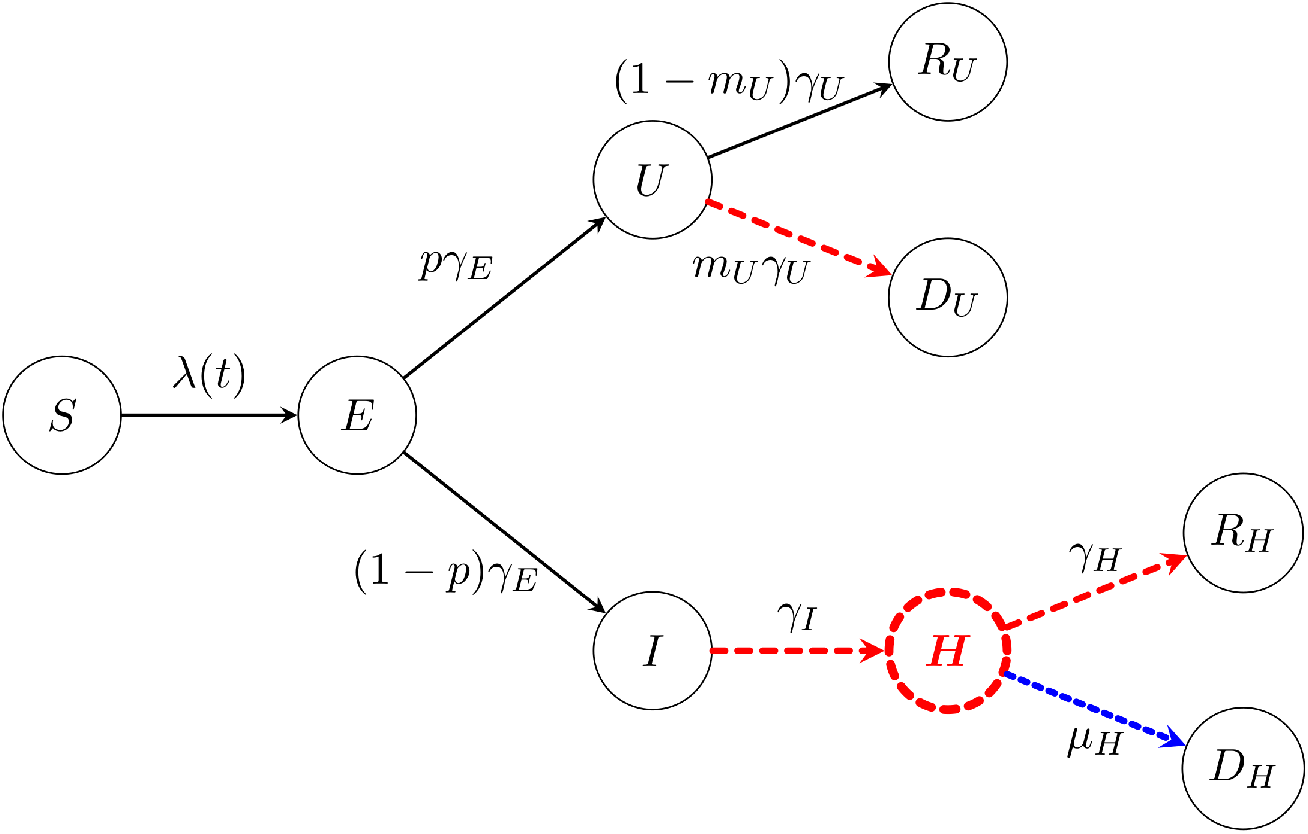
Schematic representation of the compartmental model. The susceptible population *S(t)* becomes infected through contacts with infectious individuals, *U(t)* and *I(t)*. Infected individuals incubate the disease first,^6^ and are not infectious in this state, denoted by *E(t)*, and after an incubation time, they become infectious. The compartment *U(t)* accounts for individuals that are not hospitalised; we only observe tem if they die, but not if they recover. Many models have split the *U(t)* compartment into two separate compartments (see for example Blyuss et al. (2020)^2^), one to describe individuals who are asymptomatic and the other to describe individuals who have symptoms but do not require hospitalisation. However, this approach is constrained by the lack of reliable datasets, and therefore, models of this nature rely purely on the merits of the simulations with no forecasting capabilities. For such models, it is challenging to obtain reliable data on those who are asymptomatic, especially on the scale of multiple regions/counties. Individuals in the compartment *I(t)* are eventually hospitalised and move to compartment *H(t)*. We added the hospital compartment *H(t)* into the model as a transition compartment due to the data we have access to. There are two possible outcomes for COVID-19 infections, recovery or death, denoted by *R* or *D* respectively, each subscripted with the severity of the infection. We note that there is also a difficulty gaining reliable data that considers those who are not hospitalised and recover. However, we have reliable datasets for those who die outside of hospital and thus, in the spirit of this model, are related to the not hospitalised pathway. Coloured and dashed arrows or compartments indicate that data are available: admissions to hospital (red dashed arrow from *I(t)* to *H(t)*), discharges from hospital (red dashed arrow from *H(t)* to *R*_*H*_*(t)*), daily counts of cases in hospital (red dashed *H(t)* compartment), and independent weekly data on deaths, both in hospital (red dashed arrow from *H(t)* to *D*_*H*_*(t)*), and out of the hospital settings (red dashed arrow from *U(t)* to *D*_*U*_*(t)*). A novel feature of our model and inference method is that even though information about *U(t)* is hard to come by, we can still obtain information by using the red dashed arrow between *U(t)* to *D*_*U*_*(t)*. The same thing can be said for obtaining the information on *I(t)*, by using the red dashed arrow between *I(t)* to *H(t)*. The parameters in the model regulate the rates from one compartment to the next and are described in Table 1. All parameters are inferred from the data using a minimisation process under constraints on the total population and the effective reproduction number.

The aim of our study is to propose a systematic modelling approach that addresses healthcare demand and capacity at a local level, using the Sussex datasets, by conducting healthcare demand modelling that naturally leads to a standardised framework to quantify demand generated as a result of COVID-19. This framework will facilitate short-term predictions and long-term scenario forecasting, allowing for investigations into the impact of COVID-19 on healthcare provision and planning within the local area and to mitigate long-term changes in local hospital demand as a result of further COVID-19 secondary waves. We used the local datasets collected throughout the first wave, which included local daily hospital data and weekly deaths data. Our approach differs substantially from current state-of-the-art modelling-forecasting approaches where unknown parameters driving epidemiological models have been based on various assumptions which vary substantially from one model to the other as well as variations between the domain-expertise of the researchers involved in making those assumptions. To the best of our knowledge, there is no work where the full SEIR-D model is solved and fitted to data by using only the few compartments for which data is available. This is the novelty of our approach. We impose no a priori assumptions on the values of the model parameters; instead we infer these through an inverse modelling approach by requiring the model to fit to local data in an optimal sense. From the full SEIR-D model, we derive the “observational model”, which is a representation of the full SEIR-D model described only in terms of the model parameters and compartments that are captured by the mathematical interpretation of data, in this case the observed quantities are: hospital admissions, bed occupancy, discharges, and COVID-19 related deaths (see details in the Supplementary material). In this way, by fitting the observational model to data, we obtain optimally defined values of the unknown model parameters (all the parameters shown in Figure 1), accurate to some degree of confidence.^26,27^

**Table 1:** Description of the parameters of the compartmental model, and their values when the model is fitted to the data. The values are inferred using only the data from the Sussex region, without taking any information from other regions or countries.

| Parameter | Value | Epidemiological meaning |
| --- | --- | --- |
| $\beta$ | 0.142 days<sup>-1</sup> | Average Transmission rate |
| $\gamma_E^{-1}$ | 4.67 days | Average incubation period |
| $p$ | 0.927 | Fraction of non-hospitalised infections |
| $\gamma_U^{-1}$ | 5.02 days | Average infectious period (non-hospitalised) |
| $\gamma_I^{-1}$ | 6.30 days | Average infectious period (hospitalised) |
| $\gamma_H^{-1}$ | 18.3 days | Average hospitalisation period (recovered) |
| $m_U$ | 0.0258 | Infected fatality ratio (non-hospitalised) |
| $\mu_H^{-1}$ | 16.2 days | Average hospitalisation period (deaths) |

## Methods

### Data Collection

As part of the national COVID response, all the National Health Service (NHS) hospitals in England, treating COVID-19 patients, submitted a Daily Situation Report to NHS England (NHSE). The regional data for Sussex hospitals were then sent to the Sussex Clinical Commissioning Group (CCG) who aggregated the data and combined it with the death registrations (with COVID-19 as the underlying cause of death) from the Office for National Statistics (ONS). The subsets of the hospital datasets included daily admissions, discharges, and bed occupancy. The death dataset consisted of weekly COVID-19 related deaths in hospitals and community settings (e.g. nursing homes). These datasets corresponded to the first wave of infections from 24^th^ March until 15^th^ June 2020. For the regional population count, the ONS Mid-Year Estimates (MYE) for 2018 were used. By identifying the compartments where data were available, a mathematical model was generated with the objective of forecasting local hospital demand and capacity and mortuary requirements. To mitigate changes in policy of what constituted a COVID-19 death and the procedure for recording patients with COVID-19, we account for significant levels of error n the observations. This entails that we do not explicitly distinguish between model and observational errors, but rather we compare the observations with the model solution and consider the difference to be the overall error. In particular, we are including in the error the variation due to the stochastic nature of the epidemics, since the model accounts only for the mean quantities. Although death is an absolute count, the policy regarding what constituted a COVID-19 related death changed frequently throughout the lead up to and during the lockdown period. Similarly, testing was not optimal when the hospital data collection started and so admissions and occupancy counts were retroactively edited to incorporate newly tested patients, thus the balance of total patients between days may not match up. We note that, as detailed below, our model was designed specifically to avoid the use of general testing data. In this way, we avoid dealing with the correlation between detected cases and the number of tests. The number of cases in hospitals are recorded in a systematic way in order to properly isolate the patients to avoid outbreaks and, are therefore, less dependent on the overall number of tests. In a similar manner, it is well documented that age plays an important role in the severity of a COVID-19 infection;^1,7,28,29^ however, at the beginning of the epidemic within the UK, the appropriate age-structured data simply did not exist.

### Data-driven SEIR-D modelling

The temporal dynamics of the compartmentalised epidemiological model are depicted in Figure 1, following classical approaches for formulating SIR models.^2,30^ The mathematical interpretation of the schematic diagram in Figure 1 leads to a temporal epidemiological dynamical system modelled by a system of ordinary differential equations supported by non-negative initial conditions. The full model is summarised in equations (1) – (9) in the Supplementary material.

Our model follows the general principles of SIR modelling approaches with one clear difference in that this model system is data-driven formulated where we have highlighted in dashed colour those compartments or pathways in Figure 1 where data is available within our local area. The physical justification of the SEIR-D model above is well-grounded in the modelling literature for COVID-19 and the general theory of epidemiology.^2,30^

### Inferring model parameters given hospital datasets

From the schematic diagram shown in Figure 1, we are interested in finding the optimal set of eight model parameters: *β, γ*_*E*_, *p, γ*_*U*_, *γ*_*I*_, *γ*_*H*_, *m*_*U*_, and *μ*_*H*_, such that the SEIR-D model best-fits the observed data. We estimate the parameters in the model in two steps. First, we exploit the linear relationship, arising from the mathematical model, between mortality in hospitals and discharged patients, depicted by the blue double dashed line and the red dashed line between *H(t)* and *R*_*H*_*(t)* in Figure 1, respectively, to fit the parameter *η = μ*_*H*_ *γ*_*H*_^*-1*^. The second step is to infer the remaining parameters by expressing the model in terms of the mathematical interpretation using the model parameters and compartments of the available data; we call this the observational model. Once the observational model is found, we find the maximum likelihood estimation (MLE) corresponding to the negative log-likelihood described in the Supplementary material, by means of the minimisation algorithm L-BFGS-B.^31,32^ In both cases, we explore the relationship between model parameters where we have access to reliable datasets to mitigate parameter identifiability issues.^33–35^ Details of the linear relationship of discharges and deaths in hospital and the observational model are given in the Supplementary material. We note that this two-step fitting approach is not valid in general, but the structure of the model and the data allow us to do so for this particular case, since the parameter optimized in the first step is not present in the second step. In fact, one could perform the fitting in only one step to obtain the same result. The advantage of doing the fitting in two steps is that, in the first step, we can use more appropriate techniques for the linear regression.

### Forecasting and validation

As outlined above, models of this nature often lack parameter validation and thus lack the ability to predict relatively far into the future.^8,9^ To validate the predictive power of our modified SEIR-D model (Figure 1), we used the previously outlined inference algorithm to obtain new estimates for the model parameters using only a limited number of data points, and focused on predicting the hospital admissions, discharges, and bed occupancy using a minimum of 12 and a maximum of 51 data points. This is due to the larger hospital dataset we possess, since it is recorded daily rather than recorded weekly. We evaluated the predictive power of a parameter set by performing a prediction for the next 10, 20 and 30 days, starting the day after the last data point used for the parameter estimation. By comparing the prediction with the available data, we computed the percentage of days that are correctly predicted. It was considered that a day is correctly predicted if it lies within a given tolerance of standard deviations from the available data. This approach quantifies the risk associated with the decision of selecting a certain number of days into the future, e.g. 10, 20 and 30 days. It is important to note that this approach relies on the interpretation of the data, that the data is being collected in a consistent manner, and no policy changes happen within the period of the dataset, which would incur a change in public behaviour.

## Results

### Parameter values

Using the compartmental model along with the novel inference algorithms, we derived the parameter values summarised in Table 1. Figure 2 shows the daily number of patients admitted to hospital, those in hospital and those who were discharged, respectively. To demonstrate the accuracy of the fitting procedure, we super-impose the observed datasets and their continuum mathematical counterparts as well as their 95% confidence intervals (95%CI) for these curves. It can be easily verified that the fitting captures the trends of the data and fits the majority of the data within the 95% CIs. Moreover, small perturbations in most parameters result in small changes to the overall fit of the data, whilst others result in quite a large change in the overall fit (See Supplementary material for details). This reflects how well-characterised a parameter is from the data, rather than sensitivity of the model, since the forecasting pattern is not changing significantly. Accounting for the error in the log-likelihood and the prediction technique, as well as the other sensitivity tests, demonstrates the robustness of the model when the actual data are perturbed and fitted. It is noteworthy that our set of optimal inferred parameters give a value of effective reproduction number R_t_ = 0·69 throughout the lockdown.

**Figure 2:**
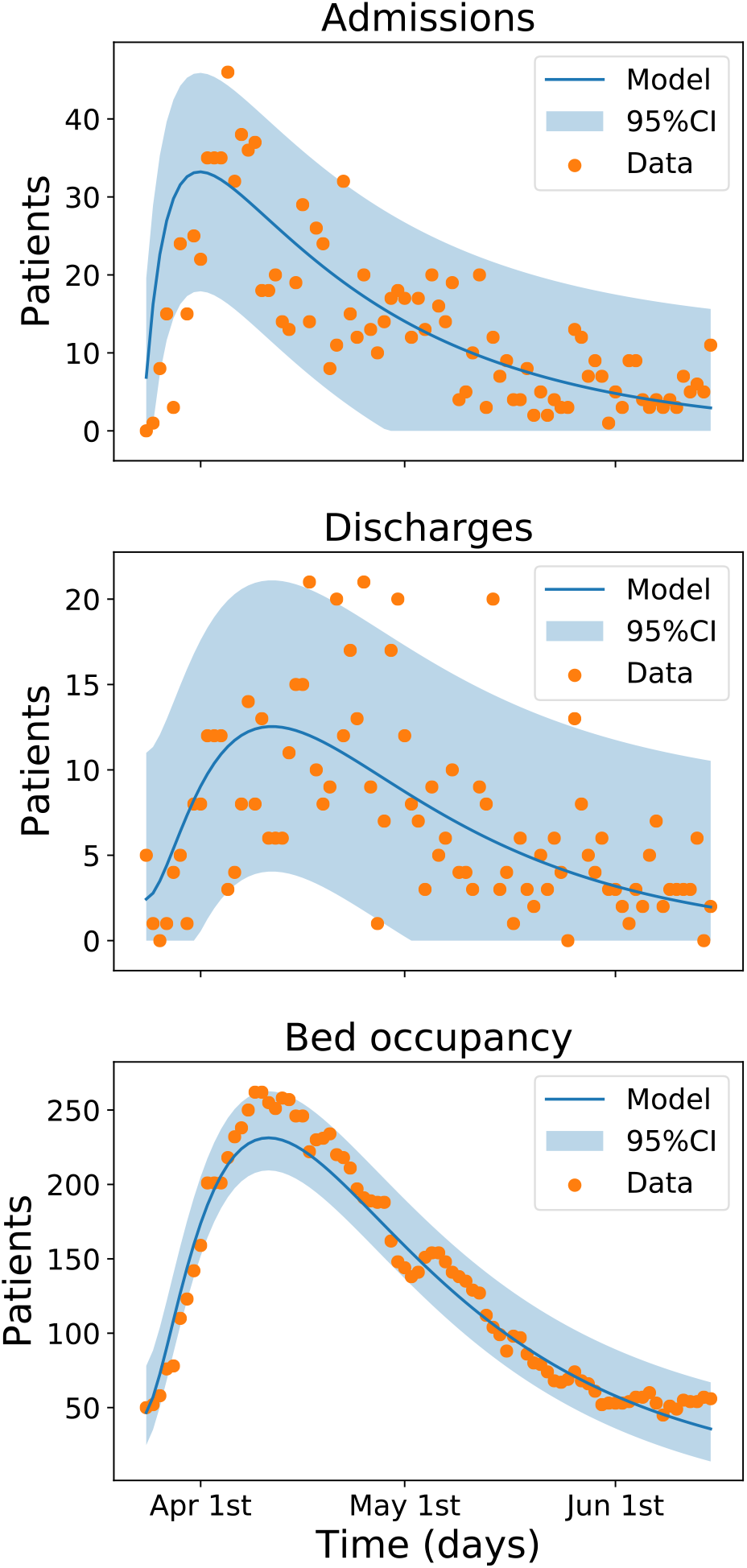
**Output of the compartmental model and comparison with data. The solid line represents the output of the model with the parameters inferred from the data. The shaded region depicts the 95% confidence interval (95%CI) computed from the data, that is, attributing all the error to measurement error. The dots correspond to observed data. Since all data are collected by manual counting and recording, there is a significant amount of noise. Furthermore, we cannot verify that the counting protocol has not changed during the period. There are between 1 and 5 outliers in each data set, out of a total of 82 data points, but generally the model captures the dynamics of the data and the situation.**

Comparisons between our parameters and those used widely in the literature are shown in Table 2.^1,6,20^ It must be noted that the physical interpretations of some of the parameters differ from one model to another, however, the overall picture appears plausible. Previous estimates of the average transmission rate and infected fatality ratio were not calibrated locally, or were based on data from other regions, for instance the Imperial College London model and other similar reports. It must be observed that the fraction of non-hospitalised cases is slightly due to its interpretation.

**Table 2:** Comparison of parameter values from different studies. There is evidence to support that one becomes infectious before presenting symptoms,^36,37^ and also that one becomes infectious after presenting symptoms.^6^ Different studies use different definitions for the incubation period, for instance time from exposure to onset of symptoms instead of time from exposure to transmissibility. This therefore has a knock-on effect on the understanding of the average infectious period. *The infected fatality ratio (IFR) in Ferguson et al. (2020)^1^ includes all cases, whilst in our model, it is limited to non-hospital infections but is heavily influenced by mortality in care-homes. **In Lourenço et al. (2020)^20^, the IFR is limited to severe infections.

| Parameter | Campillo-Funollet et al. (2020) | Ferguson et al. (2020)<sup>1</sup> | Kissler et al. (2020)<sup>6</sup> | Lourenço et al. (2020)<sup>15</sup> |
| --- | --- | --- | --- | --- |
| Average incubation period | 4·67 days | 5·1 days | 4·6 days | N/A |
| Fraction non-hospitalised cases | 0·927 | 0·956 | 0·956 | N/A |
| Average infectious period | 6·30 days | 5 days | 5 days | 4·5 days |
| Average hospitalisation | 18·3 days | 8-16 days | 6-8 days | N/A |
| Infected fatality ratio | 0·0258 | 0·009* | N/A | 0·14** |

### Predictive power of the SEIR-D model

Using the predictive power method outlined above corresponds to a total of 1776 parameters sets. The resulting values of the parameters from the inference algorithm using the subsets of data are similar to the global fit using all the available data. Figure 3 shows the results for predictions 10, 20 and 30 days into the future. To our knowledge, this is the first result of its kind that validates the forecasting in this manner.

**Figure 3:**
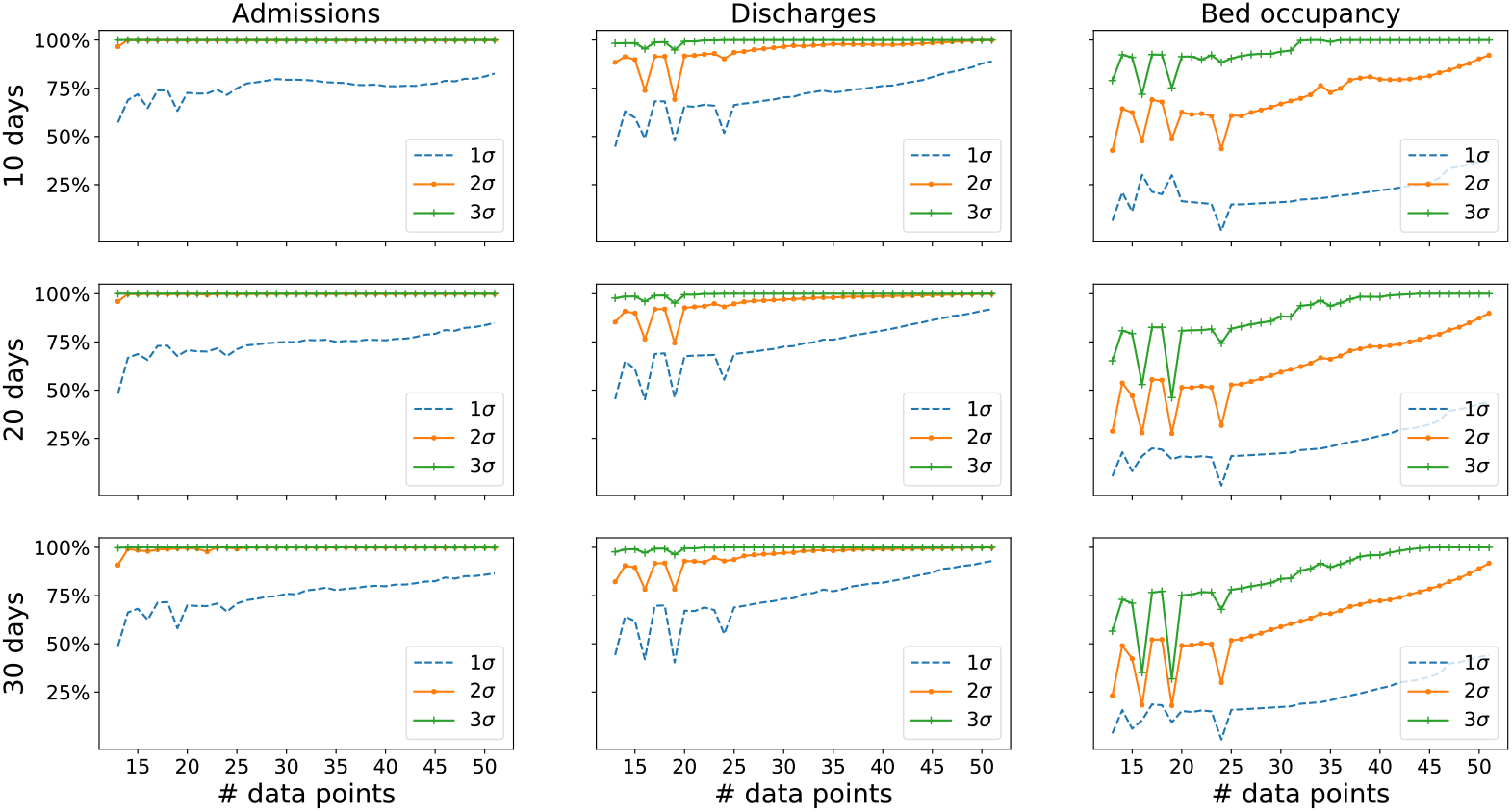
Validation of the predictive power of the model. We fitted the parameter models using all possible sequences of consecutive admissions, discharge and bed occupancy data points, from 12 to 51 points. Note that since there are only 82 data points available, we could not use more than 51 points to validate a prediction for 30 days, otherwise we will not have data to compare with. The predictive power is quantified as the number of days predicted within an accuracy of one, two, or three standard deviations of the data. There are significant differences in the predictive power for different variables. Admissions and discharges can be predicted accurately using as low as 15 data points to fit the model, whilst hospital bed occupancy requires about 30 data points to reach the same accuracy levels. Admissions/discharges and bed occupancy are different in nature: the former are rates (individuals per day), whilst the latter is an absolute count - this might explain the difference in the predictive power. In addition, bed occupancy is roughly ten times the value of admissions and discharges.

## Discussion

Predicting the local/regional resurgence of COVID-19 is the number one priority of governments and local authorities in the UK and around the world, to control and halt the local and national transmission of infection. The pandemic itself has thrown to the forefront of science the role and utility of epidemiological modelling at a time when questions of urgency, national importance, and uncertainty simultaneously come into play, thereby exposing its current limitations in terms of predictions and forecasting.^8,12^ A comment by Saltelli et al. (2020)^12^ outlines a manifesto highlighting five ways in which mathematical models should serve society. These include minding the assumptions (the minimal the better), being mindful of model complexities (hubris – balancing the usefulness of the model with the breath of its predictions), being mindful of the interests of the researchers (techniques and methodology can be limited in scope to the expertise of the researchers), being aware of the consequences (mitigate the uncertainty), and finally being mindful of the unknowns (communicating what is unknown is as important as communicating what is known). Our approach is based on these five pillars to ensure that our research outcomes are engrained and driven by reliable local surveillance data with minimal assumptions and an explicit simple data-formulated model. Predictive epidemiological modelling applied to local data has the unique ability to offer local authorities a framework for decision-making that is based on temporal trends of these local datasets. Modelling lessons learnt at the local level can possibly be transferred to the national arena to help guide data acquisition such that datasets are amenable to model-data prediction approaches as well as providing avenues for short-, medium-, and long-term forecasting.

During the early stages of COVID-19, parallels between COVID-19 and the Spanish flu (among other influenza diseases) that killed more than 50 million people with an average age of 28 years, were drawn.^1,4,6,22,38^ As a result, to mitigate and prepare for COVID-19 hospitalisations and deaths, national governments and hospitals suspended or postponed important critical diagnostic procedures/treatments, such as cancer diagnosis and treatment. Recent studies have highlighted how predictions need to be transparent and humble in order to instill confidence and invite insight and not blame.^12^ For a disease such as COVID-19, espoused wrong predictions can have a devastating effect on billions of people around the world in terms of the economy, job-security, health, education, and societal turmoil, just to mention a few. In this report, we have demonstrated that our inference process and resulting parameters allow us to produce forecasts for up to 30 days into the future to a high accuracy, for quantities of interest such as hospital bed occupancy, where such a time period can ensure that decisions, and changes in decisions, can be enacted. The underlying temporal dynamics fit the pattern of an infectious disease outbreak and does not rely solely on statistically inferred parameters,^8^ in the absence of a dynamic model. Such statistical models lack the ability for long-term forecasting. A recent review by Jewell et al. (2020)^39^ established the need for accurate forecasting in the timescales we have demonstrated to help ease public uncertainty and anxiety by aiding local policy planning in the exact manner we are using the presented results in our collaborations with the Sussex local authorities and public health departments.

It is clear from the literature that the accuracy of predictions and forecasting is closely correlated with the underlying theoretical assumptions and the use of pre-determined values of the parameters that are extracted from studies in different contexts, for instance for populations with different demographics.^8,12^ This, in turn, is driven by the lack of reliable datasets appropriate for model-data validation and sensitivity analysis. In this study, we have proposed a bottom-up approach where a model built on local datasets has the ability to guide local decision-making in terms of healthcare demand and capacity, in particular given the surge in COVID-19 secondary spikes/waves.^10,11^ Other widely used publications, such as Ferguson et al. (2020)^1^, used datasets mainly from Wuhan and other national datasets for similar infectious diseases which means that overall policy and data collection will, in general, differ to the current situation. The important highlight and applicability of our work is that we used local datasets for our modelling, and so we fully understand how the data were collected and know exactly the physical interpretation of the parameters, something that cannot be claimed by using the parameters found in the other publications of predictive modelling.^8,39^ The SEIR-D model itself is simple and transparent. Moreover, we have designed our approach in such a way that this method can be used by other regions/counties across the UK provided they have the required data, and as such, we have created a toolkit which makes our approach more accessible.^40^ This allows for users who are not familiar with mathematical modelling to use our approach and generate their own parameters to inform local policy.

Our modelling framework is not only tailored to deal with COVID-19 but can be applied to other excess death situations in summer and winter months which are known to kill thousands of people every year, provided the appropriate datasets exist. Since the framework is built around an SEIR-D model, introducing vaccinations into the model is not mathematically difficult provided we have a good understanding of the vaccination program with reliable datasets.^41–44^ Similarly, with the emergence of the new COVID-19 variants (UK, South Africa, Brazil), for example the UK VOC 202012/01 variant that emerged in the Southeast of England in November 2020, we can adapt the work by Kissler et al. (2020)^6^ to provide a multi-strain model whereby an individual either catches one strain or the other.^6,45^ Understanding the impact of these will be vitally important in the progression of dealing with the disease, however it is not clear what data will be readily available, and our observational model will need to be adapted accordingly.

Epidemic forecasting and the development of early warning systems for healthcare demand and capacity has been thrown to the forefront of epidemiological modelling. By working in close collaboration, theoreticians, local authority public health teams, and NHS planners have a unique opportunity to bring novel approaches to healthcare decision-making and planning with forecasting capabilities similar to those used for weather forecasting.

### Subsequent performance of the model post phase one of the lockdown

We continued to use the model and inference approach in the subsequent months following the lifting of COVID-19 restrictions and lockdowns. Figure 4 shows the performance of the model for hospital bed occupancy until the beginning of October 2020. The parameters of the model were re-fitted twice during the period of March to October to account for policy changes, such as implementation of lift of lockdowns and other restrictions. In these cases, the decision to re-fit was based on expert opinion, but in the future we will use model selection methods to find the optimal refitting times.

**Figure 4:**
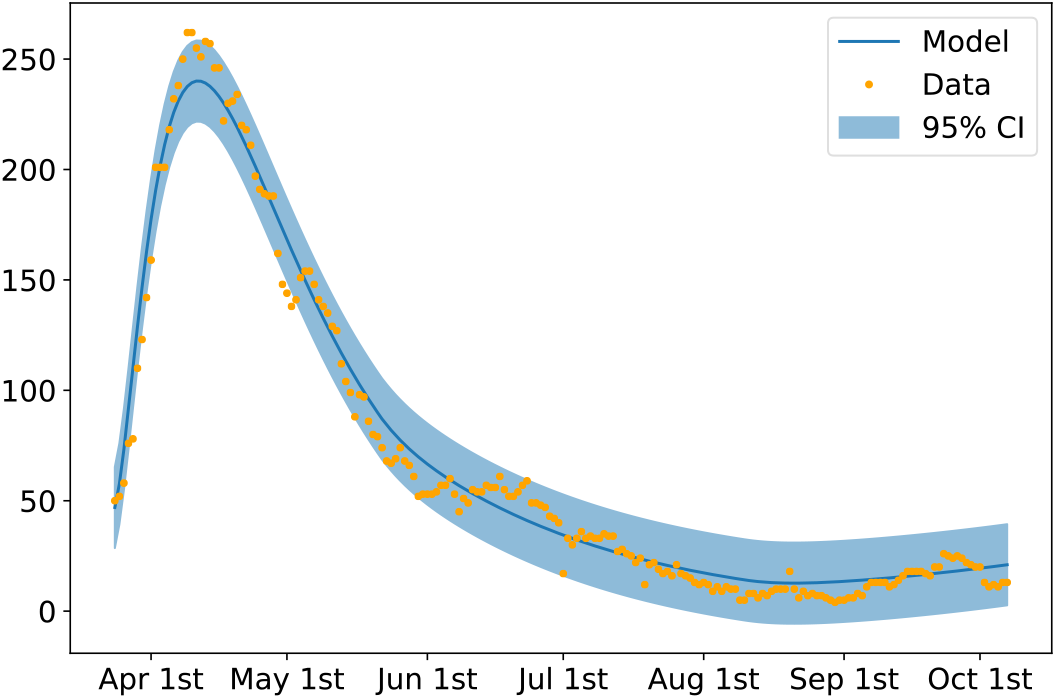
Hospital bed occupancy up to the 6^th^ October 2020. The model parameters were fitted for three different periods of time (24^th^ March to 22^nd^ May, 23^rd^ May to 10^th^ August, 11^th^ August to 6^th^ October) to reflect significant policy changes. The performance of the model is stable throughout the depicted period.

## Data Availability

All data used in the manuscript is available from public sources.

## Author Contributions

The Sussex Modelling Cell (PA, MB, WB, JC, MD, GE, KG, GP, RW, MW) conceived and presented the research questions. ECF, JVY, and AMa formulated the epidemiological model, conducted mathematical and statistical analysis and simulations of the model, and drafted the manuscript. PA and KG generated the datasets from the Sussex Clinical Commissioners and the ONS and did appropriate checks to assert the completeness and accuracy of the data. ECF and JVY assembled the figures with substantial contribution from AMa. PA, JC, MD, GE, KG, AMe and GP critically reviewed and edited the manuscript for intellectual content. All authors contributed to the final manuscript.

## Data management statement

All of the computational data output is included in the present manuscript. All images that are presented in this report are available upon request.

## Acknowledgements

JVY acknowledges postdoctoral support from the Brighton and Hove City Council, East and West Sussex County Councils.

